# Personalizing treatment selection in Crohn’s disease: a meta-analysis of individual participant data from fifteen randomized controlled trials

**DOI:** 10.1101/2023.11.10.23291837

**Authors:** Vivek A. Rudrapatna, Vignesh G. Ravindranath, Douglas V. Arneson, Arman Mosenia, Atul J. Butte, Shan Wang

## Abstract

**BACKGROUND:** Meta-analyses have found anti-TNF drugs to be the best treatment, on average, for Crohn’s disease. We performed a subgroup analysis to determine if it is possible to achieve more efficacious outcomes by individualizing treatment selection.

**METHODS:** We obtained participant-level data from 15 trials of FDA-approved treatments (N=5703). We used sequential regression and simulation to model week six disease activity as a function of drug class, demographics, and disease-related features. We performed hypothesis testing to define subgroups based on rank-ordered preferences for treatments. We queried health records from University of California Health (UCH) to estimate the impacts these models could have on practice. We computed the sample size needed to prospectively test a prediction of our models.

**RESULTS:** 45% of the participants (N=2561) showed greater efficacy with at least one drug class (anti-TNF, anti-IL-12/23, anti-integrin) over another. They were classifiable into 6 subgroups, two showing greatest efficacy with anti-TNFs (36%, N=2064). Women over 50 showed superior responses with anti-IL-12/23s. Although they represented only 2% of the trial-based cohort, 25% of Crohn’s patients at UCH are women over 50 (N=5,647), consistent with potential selection bias in trials. Moreover, 75% of biologic-exposed women over 50 did not receive an anti-IL12/23 first-line, supporting the potential value of these models. A future trial with 250 patients per arm will have 97% power to confirm the superiority of anti-IL-12/23s over anti-TNFs in these patients. A treatment recommendation tool is available at https://crohnsrx.org.

**CONCLUSIONS:** Personalizing treatment can improve outcomes in Crohn’s disease. Future work is needed to confirm these findings, and improve representativeness in Crohn’s trials.

**WHAT YOU NEED TO KNOW:** *BACKGROUND AND CONTEXT:* Patients with Crohn’s disease likely harbor different underlying susceptibilities to different treatments. Yet, clinical practice today is guided by cohort-averaging studies that ignore patient-level variation. Personalized treatment strategies are needed.

*NEW FINDINGS:* We re-analyzed data from 15 trials to model individual outcomes to different treatments. We found 6 subgroups, including women over 50 whose superior responses to anti-IL-12/23s deviate from the majority trend.

*LIMITATIONS:* This was a meta-analysis of trial data; confirmatory prospective studies are needed. Our models need to be updated to include recently approved treatments.

*CLINICAL RESEACH RELEVANCE:* Our findings confirm that heterogeneity of treatment effect does exist in Crohn’s disease. A future trial with 250 patients per arm is 97% powered to show that anti-IL-12/23s are more efficacious than anti-TNFs in women over 50. Incidentally, we also found evidence of possible selection bias into Crohn’s trials. Future work is needed to study and rectify this.

*BASIC RESEARCH RELEVANCE:* We found that patients do in fact harbor different underlying susceptibilities to different treatment mechanisms of action. But the biological basis of this is unknown. Future studies designed to elucidate this may reveal new therapeutic targets in Crohn’s disease.

## INTRODUCTION

Multiple therapies are now available for Crohn’s disease (CD), an immune disorder of the gastrointestinal tract. What remains unclear is how to select the best treatment for each patient. To date, network meta-analyses (NMAs) have been a major source of evidence on comparative efficacy and safety in CD. These studies utilize summary statistics from multiple trials to infer relative effectiveness. A recent NMA assessing three major drug classes for CD found anti-tumor necrosis factor alpha (anti-TNF) drugs to be most effective at inducing remission, followed by anti-interleukin-12/23s (anti-IL-12/23s) and anti-integrins^1^.

While NMAs have provided important information about relative effects, they have many limitations. They assume that the included trials are homogenous across multiple dimensions (e.g. cohort risk profiles, study procedures, placebo effects). They assume that the included trials are a random sample of the potential comparisons of interest (e.g. an equal chance that trials will compare drug A to B rather than A to placebo). They assume that pooled cohorts are an unbiased sample of real-world populations with active CD, justifying the application of these results to practice. Lastly, they ignore the role of patient-level variation in explaining treatment outcomes. Thus, these methods are less useful for identifying patient subgroups whose responses deviate from the majority.

Individual participant data meta-analyses (IPDMAs) are the gold-standard for meta-analyses and an alternative to NMAs^2,3^. These studies offer greater opportunities to mitigate heterogeneity across trials and to identify subgroups with different treatment responses. In a recent IPDMA performed by our group, we developed a method for normalizing the data from potentially heterogeneous clinical trials even in the absence of a consistent control group across studies^4^. We demonstrated this method, called sequential regression and simulation (SRS), in the context of nine randomized trials in CD and validated it by using those data to successfully reproduce a major secondary outcome from the recently published SEAVUE trial^5^. That trial found no average differences in effectiveness between adalimumab (anti-TNF) and ustekinumab (anti-IL-12/23).

Here we tested the hypothesis that distinct disease subgroups with different treatment responses do exist, and assessed if we can achieve more efficacious outcomes by personalizing treatment selection rather than applying general rules (e.g. recommending anti-TNF as first line for all patients without contraindications). Here, we used SRS to normalize the data from 15 trials (N=5703) corresponding to three major classes of FDA-approved drugs for Crohn’s disease (anti-TNF, anti-integrin, anti-IL-12/23). We then modeled the response to each drug class as a function of patient-level characteristics, and classified patients based on their treatment responses.

## METHODS

### DATA ACCESS

In June 2019 we queried clinicaltrials.gov to identify candidate studies (Figure 1a, Supplemental data). We confirmed 16 trials as being completed, phase 2-4, randomized, double-blinded, interventional trials of FDA-approved treatments for CD. These studies had similar cohort selection criteria and measured the Crohn’s Disease Activity Index (CDAI) at week six after treatment initiation. We successfully obtained access to the IPD for 15 studies (N=5703). These studies were conducted between 1999 and 2015 and corresponded to all six FDA-approved biologics as of 2019.

**Figure 1:**
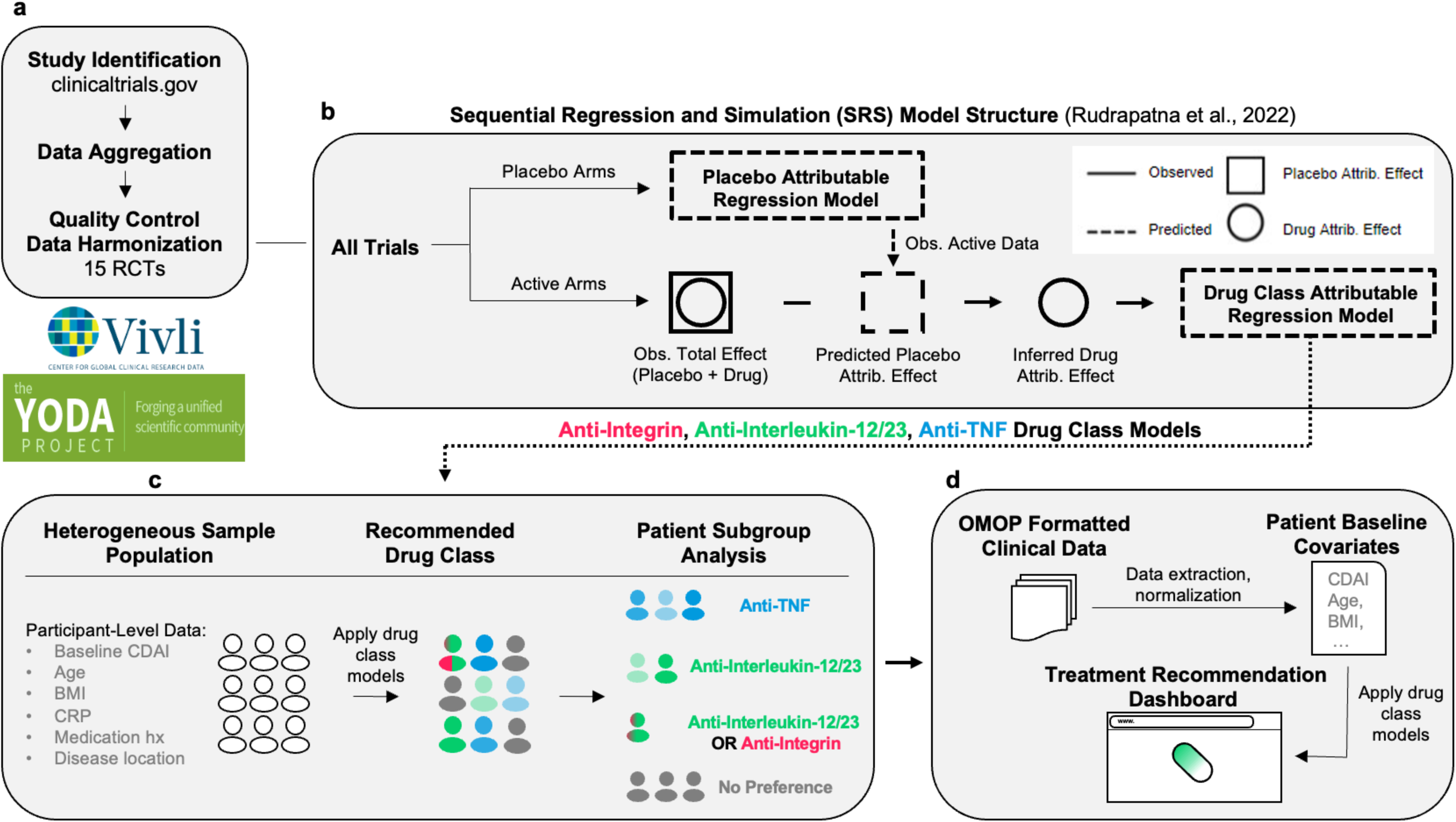
Overview. A. Clinical trials were found using clinicaltrials.gov and sought for retrieval on the YODA and Vivli platforms. Individual participant data (IPD) from trials that collected CDAI scores at week 6 visits were then aggregated and harmonised. B. Using sequential regression and simulation, a method for normalising clinical trial data against a common placebo rate, a placebo-attributable model and three drug-attributable models - anti-integrin, anti-interleukin-12/23 and anti-TNF - were developed. Disease activity reduction was partitioned into placebo attributable (square) and drug-attributable (circle) effects based on baseline covariates (age, sex, BMI, etc.). IPD (solid lines) were used to predict or simulate data (dashed lines). C. The drug-attributable models were utilised to simulate patient-level outcomes post-treatment (counterfactuals). Pairwise t-tests (p < 0.05) were conducted to compare and rank the mean responses for all drug classes - anti-integrin vs anti-interleukin-12/23, anti-integrin vs anti-TNF, and anti-interleukin-12/23 vs anti-TNF - and assign patients into one of seven subgroup memberships (see Table 3). D. Lastly, the models were re-packaged into a prototype decision support tool that uses manual inputs and optionally, OMOP-formatted data, to recommend treatments for individual patients.

### DATA CURATION

We identified nine variables that were available across all trials and thus could be used for modeling: Age, Sex, BMI, baseline CDAI, c-reactive protein (CRP), history of TNFi use, oral steroid use, immunomodulator use, and ileal involvement. Other important variables such as race and smoking were not uniformly collected across studies. We included both randomized and unblinded/open-label cohorts (Supplemental methods).

### DRUG CLASS MODELING, SUBGROUP IDENTIFICATION

We used SRS^4^ to 1) normalize all trials to a common placebo background, and 2) isolate the portion of the patient response that could specifically be attributed to a given treatment, as separate from the placebo effect (Figure 1b). For each drug class, we fit a linear mixed effects model of the drug-attributable reduction in CDAI. This outcome was modeled as a function of the above nine primary variables as fixed effects, with trial as a random effect. We compared these models to intercept-only models using the likelihood ratio test. Intercept-only models ignore the role of patient-level characteristics in determining treatment responses and reflect the assumptions of NMAs.

We applied the three fitted models to each of the 5703 participants to simulate their response under each of three counterfactual scenarios: treatment with an anti-TNF vs anti-integrin vs anti-IL-12/23 (Figure 1c). The inferred normal distributions of the conditional mean responses were pairwise compared using the medians and standard errors of the bootstrapped predictions (Supplemental Methods). We used a p=0.05 threshold to identify patients belonging to a subgroup, defined as a distinct pattern of ordinal preferences across all three drug classes.

### WEB APPLICATION

We prototyped a decision support tool (https://crohnsrx.org). This application utilizes manually-inputted data to produce recommendations. For users seeking to deploy this dashboard locally, an additional mode that directly sources inputs from OMOP-formatted databases is available (Figure 1d).

## RESULTS

### COHORT CHARACTERISTICS

Our cohort consisted of 5703 participants, drawn from fifteen trials of all FDA-approved biologics as of 2019. These biologics corresponded to three drug classes: anti-TNFs, anti-IL-12/23s, and anti-integrins. The members of our cohort were generally similar by their univariate characteristics across trials (Table 1).

**Table 1:**
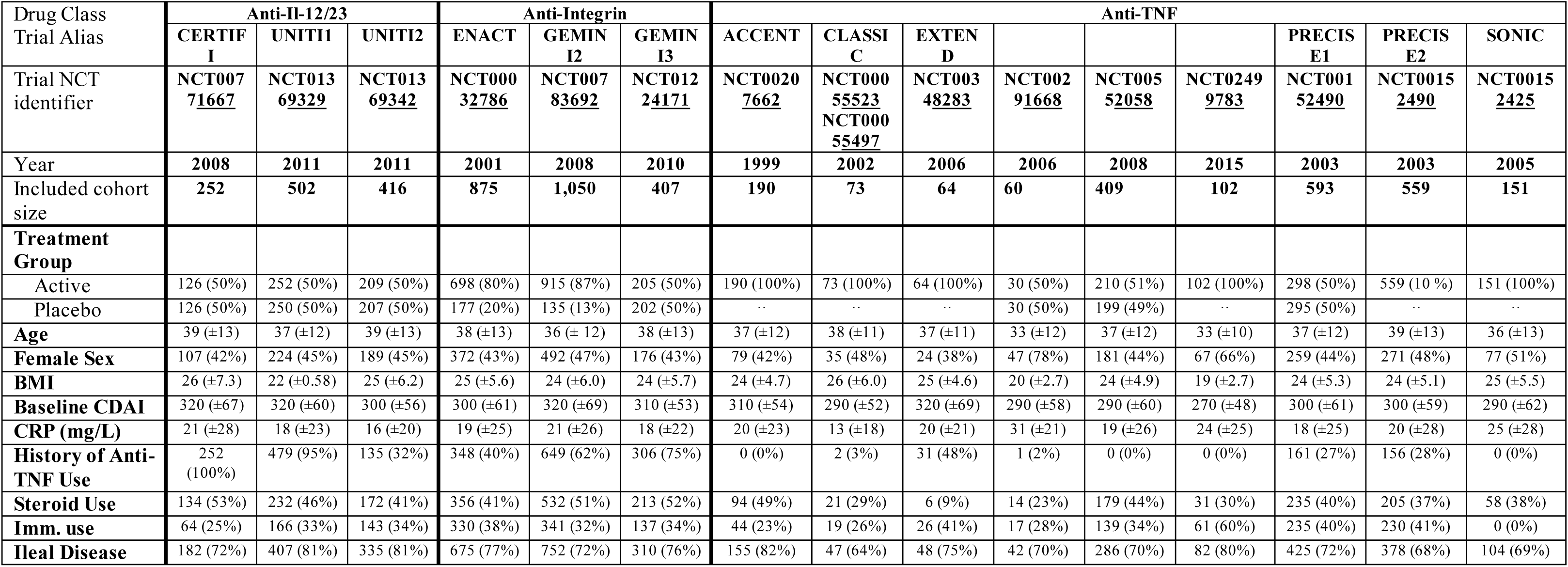
Baseline characteristics of the meta-analyzed study cohort, stratified by trial. Characterization of baseline covariates of included studies. Placebo arms from ACCENT, CLASSIC, EXTEND, NCT02499783, PRECISE2, and SONIC studies were not included due to the absence of a 6-week parallel arm placebo group (see Supplementary Figure 1). Continuous variables are reported as mean (standard deviation) and binary variables are reported as count (proportion). CRP = c-reactive protein, TNF = tumor necrosis factor.

### PLACEBO MODEL

To address the potential bias that could result from a naive pooling of subjects across trials, we used sequential regression and simulation to normalize the data and analytically separate the drug-attributable component of the patient response from the placebo effect^4^.

Using the subset of participants assigned to placebo (N=1621), we modeled their week 6 response as a function of all captured covariates and study year (fixed effects) as well as trial of origin (random effect). This model was significant (p<0.001; Table 2). We identified six predictors of the placebo effect. The coefficient for study year was negative, suggesting a reduction in measured placebo effects over time. Male sex was associated with 27 points less of a placebo effect. Baseline CDAI was also a significant predictor: every 100 points of a higher baseline CDAI (restricted by trial eligibility criteria to fall between 220-450) was associated with 33 points more of spontaneous improvement after 6 weeks. This was consistent with regression to the mean. Age and CRP were also significant albeit with small estimated effects. Most of the explainable variation in the placebo effect was accounted for by these explicitly captured clinical factors and study year; only 1% of the total variation was attributable to other non-specific heterogeneity across the included trials.

**Table 2:** Linear mixed effect regression models of the reduction in CDAI at week 6. We fit a total of four linear mixed effects regression models: one placebo model and three nested models of the drug class-attributable response. Rows correspond to the fixed effect parameters of each model, and columns correspond to the estimated coefficients, standard errors, and Wald test p-values with bolding corresponding to significance at the 0·05 level. Year was not used for the drug class models due to insufficient variation (few trials per drug class, clustered together in calendar time).

| Predictors | Placebo |  |  | Anti-IL-12/23 |  |  | Anti-Integrin |  |  | Anti-TNF |  |  |
| --- | --- | --- | --- | --- | --- | --- | --- | --- | --- | --- | --- | --- |
|  | *Estimate* | *Std. Error* | *p* | *Estimate* | *Std. Error* | *p* | *Estimate* | *Std. Error* | *p* | *Estimate* | *Std. Error* | *p* |
| Intercept | 74.69 | 9.48 | **<0.001** | 22.19 | 22.01 | 0.314 | 36.82 | 7.34 | **<0.001** | 54.96 | 9.80 | **<0.001** |
| Year (Centered) | -1.96 | 0.99 | **0.048** | .. | .. | .. | .. | .. | .. | .. | .. | .. |
| Baseline CDAI (Centered) | 0.33 | 0.04 | **<0.001** | -0.03 | 0.06 | 0.640 | 0.02 | 0.04 | 0.590 | 0.11 | 0.04 | **0.002** |
| Age (Centered) | 0.41 | 0.18 | **0.025** | 0.30 | 0.30 | 0.313 | -0.54 | 0.19 | **0.004** | -1.23 | 0.18 | **<0.001** |
| BMI (Centered) | 0.72 | 0.43 | 0.098 | -1.69 | 0.75 | **0.024** | -0.35 | 0.39 | 0.380 | -0.19 | 0.44 | 0.660 |
| CRP (mg/L) (Centered) | -0.22 | 0.10 | **0.022** | 0.48 | 0.16 | **0.002** | 0.12 | 0.09 | 0.196 | 0.35 | 0.08 | **<0.001** |
| Sex: Male | -27.31 | 5.24 | **<0.001** | 28.00 | 12.06 | **0.021** | 1.66 | 4.49 | 0.712 | 7.03 | 6.09 | 0.249 |
| History of Anti-TNF Use | 2.22 | 4.42 | 0.616 | -6.41 | 7.74 | 0.408 | -4.03 | 4.32 | 0.351 | 0.68 | 4.16 | 0.871 |
| Steroid Use | 0.32 | 4.44 | 0.943 | 19.82 | 7.60 | **0.009** | 4.97 | 4.32 | 0.251 | -1.69 | 4.30 | 0.694 |
| Immunomod. Use | -1.74 | 4.69 | 0.711 | 0.54 | 8.28 | 0.948 | -4.35 | 4.54 | 0.338 | -1.70 | 4.55 | 0.708 |
| Ileal Disease | 4.88 | 5.05 | 0.333 | -9.14 | 9.64 | 0.344 | -8.15 | 4.97 | 0.102 | -7.29 | 4.58 | 0.111 |

### DRUG CLASS MODELS

We used the placebo model to calculate the mean placebo-attributable response for each participant assigned to receive active treatment (N=4082) and subtract this from their observed response, leaving behind the drug-attributable reduction in CDAI. We then used the residuals to fit three additional mixed effects models, one per drug class.

The drug class models were significant (p<0.01 for all; Table 2). We identified 10 predictors across drug classes. Efficacious responses to IL12/23s were positively associated with male sex (28 additional points of CDAI reduction) and steroid use (20 additional points). Elevated CRP was associated with a positive response to IL12/23s, whereas elevated BMI was associated with a negative response. For the anti-integrin class, each decade of life was associated with 5 points less of a response on the CDAI. Lastly, for the anti-TNF class we identified 3 additional predictors of efficacy beyond the intercept term. Elevations in baseline CDAI and CRP were associated with increased efficacy, whereas age was inversely associated (12 points less of CDAI reduction for each decade).

To improve the efficiency of future trials, we compared significant coefficients identified in the placebo and active treatment models. Five coefficients had opposite effects: age, BMI, CRP, male gender, and ileal involvement (Table 2). These results implied that young males with lower BMIs, elevated CRP and colonic disease would be expected to have the widest margin of difference between placebo and treatment arms, and thus the greatest power to detect evidence of efficacy. This finding also underscored the importance of separating placebo- and drug-attributable effects using separate regression models; a regression model lacking these implied interaction terms would miss these findings.

### SUBGROUPS

We simulated potential outcomes for all participants under each drug class and performed pairwise t-tests to rank-order treatment preferences and define subgroups. We identified six subgroups (Table 3) in our primary analysis, and three more when using a less stringent p-value (Supplemental Table 2). Most participants (55%, N=3142) did not show strong evidence for superior efficacy with any one class over another. Most of the others showed evidence for an anti-TNF being best or tied-for-best (42%, N=2418). These results explain prior findings favoring anti-TNFs as being the result of using ‘majority vote’ statistical methods in a situation where most participants ‘abstain.’

**Table 3:**
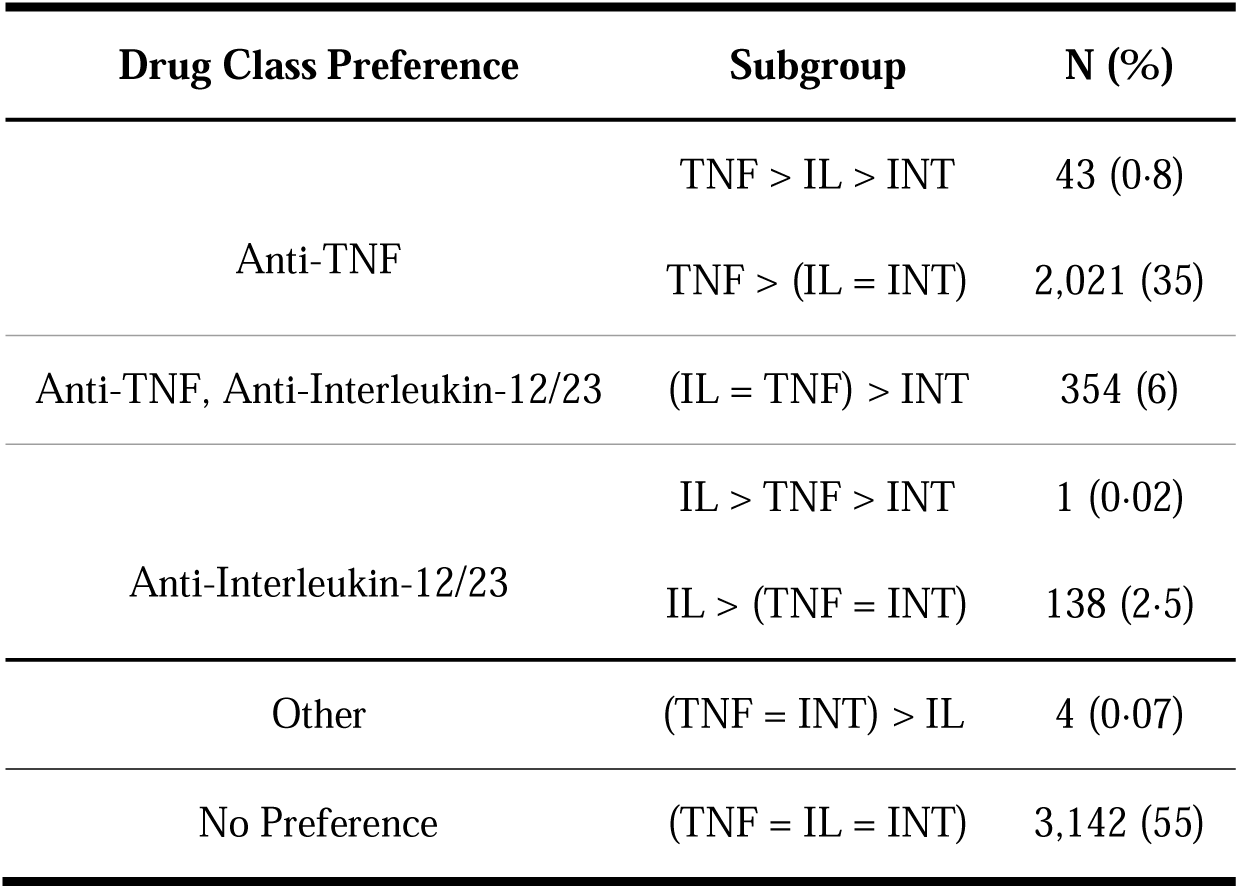
Treatment subgroups. The finalized mixed effects models were used to simulate counterfactual outcomes under all possible treatment scenarios. The modeled outcomes and the associated uncertainties in these outcomes were used to perform pairwise t-testing to assess evidence for rank-ordered preferences across drug classes. Distinct patterns of rank-orderings were used to establish membership in one of 6 subgroups. Subjects without sufficient statistical evidence (alpha = .05) of a more efficacious response to any one drug classes were placed into a 7^th^ category (no preference). TNF = anti-tumor necrosis factor, IL = anti-interleukin-12/23, INT = anti-integrin.

| Drug Class Preference | Subgroup | N (%) |
| --- | --- | --- |
| Anti-TNF | TNF > IL > INT | 43 (0.8) |
|  | TNF > (IL = INT) | 2,021 (35) |
| Anti-TNF, Anti-Interleukin-12/23 | (IL = TNF) > INT | 354 (6) |
| Anti-Interleukin-12/23 | IL > TNF > INT | 1 (0.02) |
|  | IL > (TNF = INT) | 138 (2.5) |
| Other | (TNF = INT) > IL | 4 (0.07) |
| No Preference | (TNF = IL = INT) | 3,142 (55) |

However, 139 participants showed evidence of superior efficacy with an anti-IL-12/23, achieving 40 points greater reduction on the CDAI compared to the other drug classes (Figure 2; Supplementary Table 3). 50% of these patients were predicted as achieving clinical response (CDAI reduction of 100 points or more) at week 6, compared to only 3% with an anti-TNF. This subgroup was predominantly female, over the age of 50, had a history of anti-TNF exposure, had relatively lower CDAIs at baseline, and were receiving steroids. This subgroup corresponded to only 2% of the trial population.

**Figure 2:**
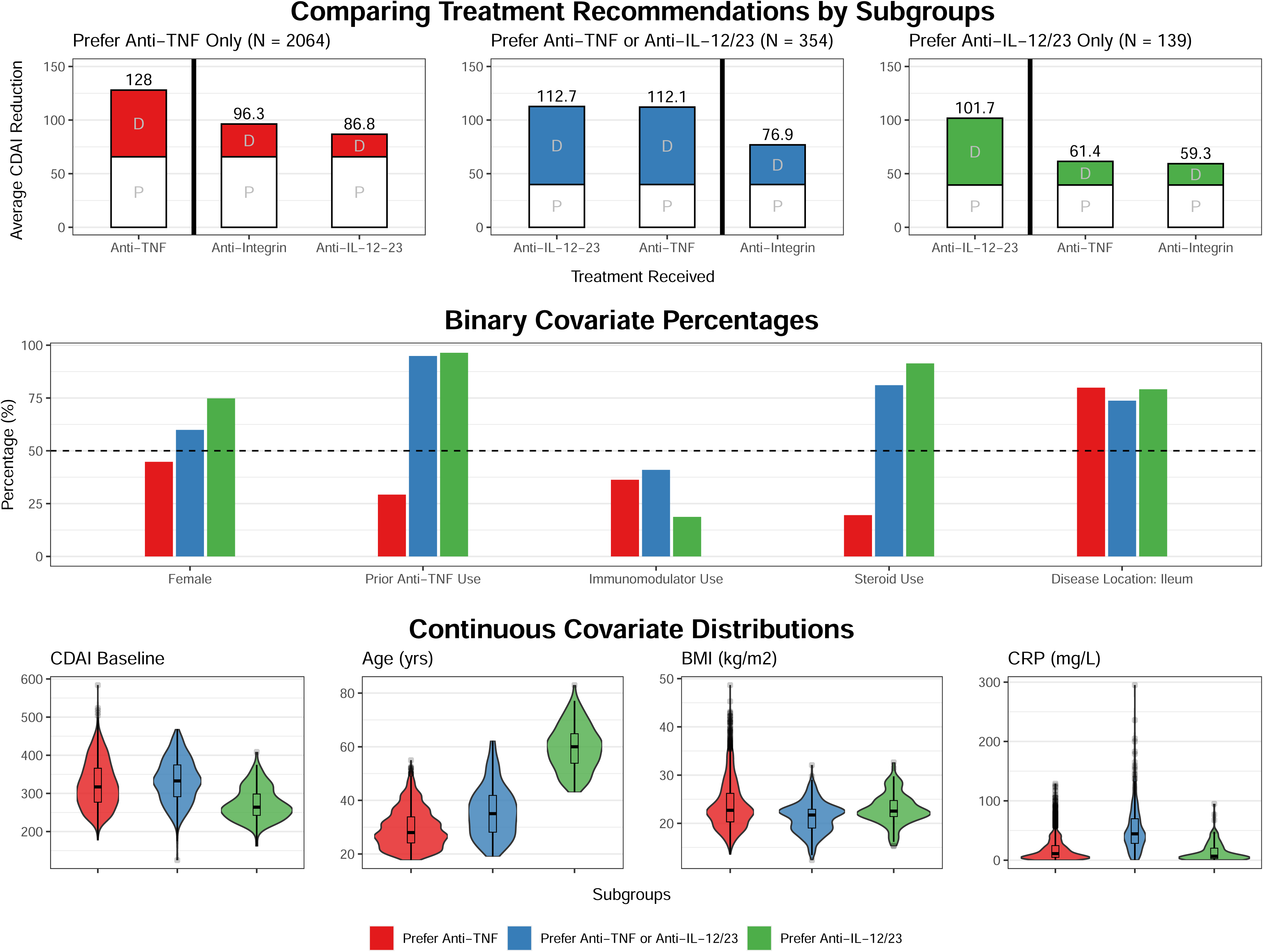
Subgroup analysis. Detailed comparison of three major subgroup cohorts found in the trial-based cohort (N=5703): prefer anti-TNF only (N = 2061, red), prefer anti-TNF or anti-IL-12/23 (N = 355, blue), and prefer anti-IL-12/23 only (N = 139, green). A. The bar plots on the left show the average placebo (P) and drug-class (D) attributable effects for each subgroup. Superior drug classes (left of bolded vertical line) reduce disease activity (CDAI reduction) by 30-40 points more on average compared to non-superior drug-classes (right of bolded vertical line). B. The plots on the right compare the proportions and distributions of covariates for each subgroup.

Given this, we wondered if a decision support tool might have any measurable value in clinical practice, compared to an easier-to-remember strategy of recommending an anti-TNF to any patient lacking a contraindication. We queried the University of California Health Data Warehouse, a multicenter database of health records data, to identify potential patients who might belong to this subgroup and thus could benefit from a personalized treatment recommendation tool. We found that 25% of the patients seen for Crohn’s disease were women over the age of 50 (N=5,647; 2012-2022) (Supplementary Figure 5). This striking difference in cohort prevalence (25% at the University of California vs 2% in the trials) suggested the possibility of implicit selection bias in these trials. Supporting this view, we found Black participants to be significantly underrepresented (2% in the trials; Supplementary Table 4).

When limiting our queries to the timeframe when all drug classes were FDA-approved, we noted that 75% of biologic-exposed women over 50 did not receive an anti-IL12/23 first-line. This suggested a potential future role for software-aided treatment optimization.

Since the existence of this anti-IL-12/23-preferring subgroup was a new and potentially testable hypothesis raised by this analysis, we performed a sample size calculation to determine the feasibility of verifying this in a prospective study (Supplementary Table 5). We calculated that a trial with 250 participants in each arm would have 87% power to show superiority of anti-IL-12/23s over anti-TNFs in all patients over the age of 50. If further restricted to just women over 50, this potential trial was calculated as having 97% power.

### DECISION SUPPORT

To bridge these findings to the clinic we have prototyped a decision support tool (https://crohnsrx.org). It uses manual inputs on patient-level features to produce treatment recommendations (Figure 3). We have provided additional guidance to help clinicians interpret the output and avoid incorrectly using the tool on patients who do not resemble the subjects used to train the model.

**Figure 3:**
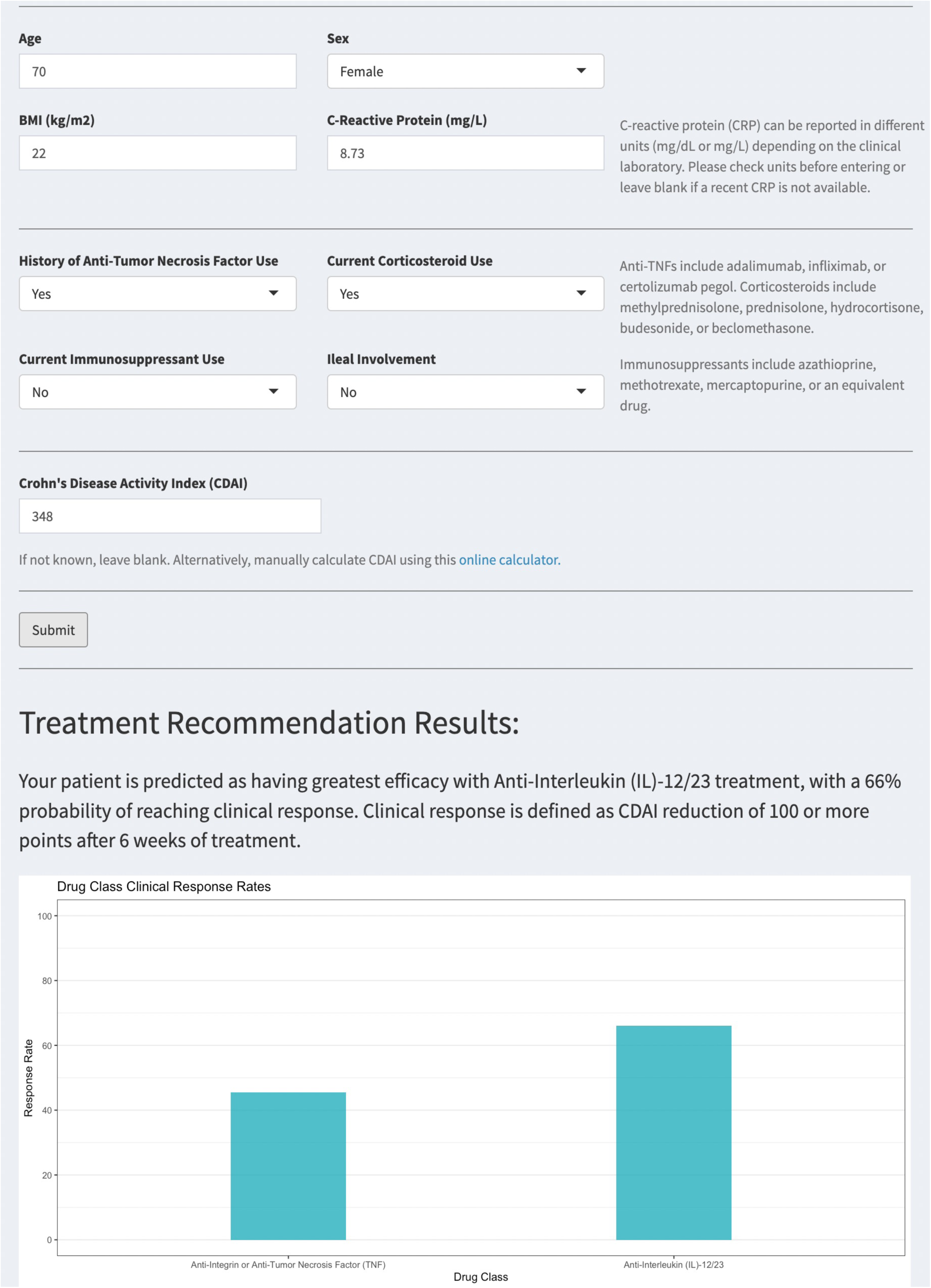
Treatment recommendation dashboard. Example user interface and output of R Shiny treatment recommendation dashboard.

## DISCUSSION

We performed a subgroup analysis of individual participant data from 15 trials of treatments for Crohn’s disease (CD). Our primary findings can be summarized as follows: 1) patients with CD likely harbor different underlying preferences towards different treatments, and these preferences are partially predictable using clinical features, 2) most trial participants do not appear to have superior efficacy with anti-TNFs drugs, a potentially unexpected finding given prior literature, 3) there appears to be evidence of significant implicit selection bias into registrational trials, and 4) the use of statistically-based decision support tools may improve patient outcomes in clinical practice. Secondary results include 1) newly-identified features that predict patient-level responses to different drugs as well as placebo, and insights as to how they could be used to design more efficient clinical trials, and 2) sample size calculations supporting the feasibility of testing this model’s predictions prospectively. These findings add to a growing body of evidence that shifts away from a ‘one-size-fits-all’ treatment paradigm and towards precision medicine.

Our work builds on previous evidence synthesis efforts in CD, particularly NMAs^1,6^. The most recent of these found that anti-TNF drugs appear to be the most efficacious drug for inducing clinical remission^1^. Although we used a similar set of trials as that study, we came to a slightly different conclusion: most of the subjects in these trials *do not* appear to preferentially benefit from any of three currently approved drug classes. Instead, we found that patients favoring anti-TNFs were actually in the minority, albeit a large one (42%). This apparent contradiction can be understood as the result of an “ecological fallacy”, where one incorrectly deduces that a cohort-averaged effect also applies to each member of the cohort. An apt analogy would be of an election where the majority abstains, and the next largest constituency ‘votes’ for an anti-TNF.

Thus, our findings are in fact consistent with prior NMAs that instead rely on aggregate statistics from trials. However, these findings more generally suggest that the field of evidence synthesis must increasingly embrace IPD to generate results that are more precise and less susceptible to misinterpretation. Methods such as SRS can add additional credibility and reduce the dependence on strong homogeneity assumptions implicit in traditional pooled analyses of IPD. This method also enables deeper insights into the overall patient response as the result of two distinguishable effects: placebo-attributable and drug-attributable. The predictors of the placebo effect that we identified here were consistent with the prior literature^7^. Yet we identified more predictors than have previously been reported, likely because our method implicitly accounts for drug-by-effect interactions that are often unmodeled in one-step IPD meta-analyses^2,3^. The value of these findings, beyond that of scientific interest into how clinical features reflect treatment susceptibility, is also practical. Our results suggest ways to design clinical trials with greater power.

Another important but unexpected finding is evidence of significant selection bias in Crohn’s RCTs. Some degree of selection bias is to be expected of all trials insofar as they contain additional inclusion and exclusion criteria that are not a requirement for receiving clinical care. Indeed, we and others have observed this in the context of comparing RCT- and real-world cohorts^8–10^. What has been unclear, however, is the extent to which these biases may distort treatment outcomes. We identified a subgroup of anti-IL-12/23-preferring patients, mostly women over 50, that represented a miniscule fraction of trial subjects (2%). Yet, the typical prevalence of these patients as seen across 6 medical centers at the University of California suggests that as many as 25% patients fall into this demographic. Of course, gender, older age, and race are not explicit exclusionary criteria in these registrational trials. Thus, it appears that these patients are systematically being under-enrolled. Future studies are needed to determine if this is the result of patient preferences, provider biases, or other factors.

Artificial intelligence is playing a growing role in healthcare, with a proliferation of data-informed software tools that can help clinicians make better decisions. Yet, many uncertainties remain as to how these models should be tested before they can be trusted and deployed in practice^11^. Prospectively testing our model in its entirety is generally a challenge because it makes predictions across the continuum of patient features. Many patients did not appear to have a strong treatment preference, making it challenging to envision a study that ‘confirms’ this null hypothesis. By contrast, the aforementioned subgroup of older women is ideal from a prospective standpoint: 1) it is a scientifically new finding, 2) it demonstrates a treatment effect that can be evaluated via null hypothesis testing, and 3) its members are sufficiently prevalent in real-world settings, increasing the potential feasibility of subject recruitment.

Separate from testing the statistical model is testing the decision support prototype itself. We have released an early version (https://crohnsrx.org) to facilitate early user feedback and enhancement. Future work is needed to develop this into an EHR-embedded tool that supports seamless, timely, and trustworthy recommendations at the point of care.

Strengths of this work include the strength and quality of the underlying data, the use of the SRS method to address bias and reveal new insights, as well as several important findings impacting the science, study, and care of patients with Crohn’s disease.

We acknowledge several limitations. This was a post-hoc analysis of randomized trials, and we cannot completely exclude residual biases. Prospective studies are needed to test these findings, particularly given apparent selection biases that could degrade the application of trial-based insights to clinical practice. There were several variables that we wanted to include in our models such as race/ethnicity and smoking. Unfortunately, these data were not well-captured across the trials. We found a large amount of unexplained variability in patient outcomes, roughly 90%. This in and of itself is not a weakness of our study, but rather reflects the large scope of future work that is needed to explain patient outcomes in Crohn’s disease. Some of this work will require discovering new biomarkers (e.g., metagenomic, metabolomic), none of which were available to us. Other work will be needed to increase the objectivity of disease activity measures in Crohn’s disease and reduce unwanted variation.

In conclusion, we performed an IPD meta-analysis of RCTs in Crohn’s disease. We identified multiple subgroups with different preferential responses to different drug classes, including one subgroup of women over 50 who may respond favorably to anti-IL-12/23s. We uncovered potential evidence of selection bias into clinical trials and suggest ways to improve the efficiency and equity of these gold-standard studies. Lastly, we developed a prototype decision support tool that implements these findings, in the hopes that it will help improve treatment selection and patient outcomes in Crohn’s disease.

## Data Availability

The raw data are owned by the sponsors of the trials that were meta-analyzed in this work. This data may be accessed for reproduction and extension of this work following an application on the YODA and Vivli platforms and execution of a data use agreement. The analytical code is available at https://github.com/rwelab/CrohnsRx.

https://vivli.org

https://yoda.yale.edu

## ACKNOWLEDGEMENTS

The authors thank all the trial sponsors (Johnson & Johnson, AbbVie, UCB, Takeda, Biogen) for contributing the necessary participant-level data to carry out this study, and the participants of these studies consenting for their deidentified data to be shared in this way. They thank both the Yale Open Data Access Project and Vivli for enabling data access on a single platform. They thank Isabel Elaine Allen, Chiung-Yu Huang, and Mi-Ok Kim for biostatistical advice. They thank Cong Zhou for her contributions to earlier versions of this work.

This study, carried out under YODA Project #2018-3476, used data obtained from the Yale University Open Data Access Project, which has an agreement with Janssen Research & Development LLC. The interpretation and reporting of research using this data are solely the responsibility of the authors and does not necessarily represent the official views of the Yale University Open Data Access Project or Janssen Research & Development LLC.

This publication is based on research using data from data contributors Johnson & Johnson, AbbVie, UCB, Takeda, Biogen that has been made available through Vivli, Inc. Vivli has not contributed to or approved, and is not in any way responsible for, the contents of this publication.

The authors thank the Center for Data-driven Insights and Innovation at UC Health (CDI2; https://www.ucop.edu/uc-health/functions/center-for-data-driven-insights-and-innovationscdi2.html), for its analytical and technical support related to use of the UC Health Data Warehouse and related data assets.

## FUNDING

Research reported in this publication was supported by the National Library of Medicine of the National Institutes of Health under Award Number K99LM014099, the National Institute of Diabetes and Digestive and Kidney Diseases under Award Number 5T32DK007007-48 the National Center for Advancing Translational Sciences, National Institutes of Health, through UCSF-CTSI Grant Number UL1 TR001872. Its contents are solely the responsibility of the authors and do not necessarily represent the official views of the NIH. Additional funding support for data access was provided by the University of San Francisco (USF). The contents of this manuscript are solely the responsibility of the authors and do not necessarily represent the official views of the NIH, UCSF, or USF. The funders had no role in the study design, the collection, analysis, and interpretation of data, in the writing of the report, in the decision to submit the article for publication. The authors confirm their independence from the funders. All authors, external and internal, had full access to all of the data (including statistical reports and tables) in the study and can take responsibility for the integrity of the data and the accuracy of the data analysis.

## AUTHOR CONTRIBUTIONS

VAR conceived the study and obtained access to the data. SW, VGR, DVA, and VAR designed the study, analysed the data. AM performed the risk of bias assessment. SW performed an independent review of the analytical code. VAR wrote the first draft of the manuscript. All authors interpreted the data and critically edited the manuscript.

## POTENTIAL COMPETING INTERESTS

VAR received grant support from Janssen, Alnylam, Merck, Genentech, Takeda, and Blueprint Medicines for unrelated work during this study. DVA is currently an employee at Bristol Myers Squibb. AJB is a co-founder and consultant to Personalis and NuMedii; consultant to Mango Tree Corporation, and in the recent past, Samsung, 10x Genomics, Helix, Pathway Genomics, and Verinata (Illumina); has served on paid advisory panels or boards for Geisinger Health, Regenstrief Institute, Gerson Lehman Group, AlphaSights, Covance, Novartis, Genentech, and Merck, and Roche; is a shareholder in Personalis and NuMedii; is a minor shareholder in Apple, Meta (Facebook), Alphabet (Google), Microsoft, Amazon, Snap, 10x Genomics, Illumina, Regeneron, Sanofi, Pfizer, Royalty Pharma, Moderna, Sutro, Doximity, BioNtech, Invitae, Pacific Biosciences, Editas Medicine, Nuna Health, Assay Depot, and Vet24seven, and several other non-health related companies and mutual funds; and has received honoraria and travel reimbursement for invited talks from Johnson and Johnson, Roche, Genentech, Pfizer, Merck, Lilly, Takeda, Varian, Mars, Siemens, Optum, Abbott, Celgene, AstraZeneca, AbbVie, Westat, and many academic institutions, medical or disease specific foundations and associations, and health systems. Atul Butte receives royalty payments through Stanford University, for several patents and other disclosures licensed to NuMedii and Personalis. Atul Butte’s research has been funded by NIH, Peraton (as the prime on an NIH contract), Genentech, Johnson and Johnson, FDA, Robert Wood Johnson Foundation, Leon Lowenstein Foundation, Intervalien Foundation, Priscilla Chan and Mark Zuckerberg, the Barbara and Gerson Bakar Foundation, and in the recent past, the March of Dimes, Juvenile Diabetes Research Foundation, California Governor’s Office of Planning and Research, California Institute for Regenerative Medicine, L’Oreal, and Progenity. The authors have declared that no actual competing interests exist.

## DATA SHARING PLAN

The raw data are owned by the aforementioned trial sponsors. The data may be accessed for reproduction and extension of this work following an application on the YODA and Vivli platforms and execution of a data use agreement. The analytical code is available as supplemental data files accompanying this manuscript.

## Notes

### Competing Interest Statement

VAR received grant support from Janssen, Alnylam, Merck, Genentech, and Blueprint Medicines for unrelated work during this study. DVA is currently an employee at Bristol Myers Squibb. AJB is a co-founder and consultant to Personalis and NuMedii; consultant to Mango Tree Corporation, and in the recent past, Samsung, 10x Genomics, Helix, Pathway Genomics, and Verinata (Illumina); has served on paid advisory panels or boards for Geisinger Health, Regenstrief Institute, Gerson Lehman Group, AlphaSights, Covance, Novartis, Genentech, and Merck, and Roche; is a shareholder in Personalis and NuMedii; is a minor shareholder in Apple, Meta (Facebook), Alphabet (Google), Microsoft, Amazon, Snap, 10x Genomics, Illumina, Regeneron, Sanofi, Pfizer, Royalty Pharma, Moderna, Sutro, Doximity, BioNtech, Invitae, Pacific Biosciences, Editas Medicine, Nuna Health, Assay Depot, and Vet24seven, and several other non-health related companies and mutual funds; and has received honoraria and travel reimbursement for invited talks from Johnson and Johnson, Roche, Genentech, Pfizer, Merck, Lilly, Takeda, Varian, Mars, Siemens, Optum, Abbott, Celgene, AstraZeneca, AbbVie, Westat, and many academic institutions, medical or disease specific foundations and associations, and health systems. Atul Butte receives royalty payments through Stanford University, for several patents and other disclosures licensed to NuMedii and Personalis. Atul Butte's research has been funded by NIH, Peraton (as the prime on an NIH contract), Genentech, Johnson and Johnson, FDA, Robert Wood Johnson Foundation, Leon Lowenstein Foundation, Intervalien Foundation, Priscilla Chan and Mark Zuckerberg, the Barbara and Gerson Bakar Foundation, and in the recent past, the March of Dimes, Juvenile Diabetes Research Foundation, California Governor's Office of Planning and Research, California Institute for Regenerative Medicine, L'Oreal, and Progenity. The authors have declared that no actual competing interests exist.

### Clinical Protocols

https://www.crd.york.ac.uk/PROSPERO/display_record.php?RecordID=157827

https://yoda.yale.edu/sites/default/files/files/YODA%20Project%20Protocol%202018-3476%20-%2019-07-01.pdf

### Funding Statement

This publication was supported by the National Center for Advancing Translational Sciences, National Institutes of Health, through UCSF-CTSI Grant Number UL1 TR001872. Its contents are solely the responsibility of the authors and do not necessarily represent the official views of the NIH. VAR was partially supported by an institutional training grant (5T32DK007007-48) funded by the NIH and administered by UCSF. Additional funding support for data access was provided by the University of San Francisco.

### Author Declarations

This study was approved by the UCSF Institutional Review Board (#18-24588)

