## Supplemental Material for "Personalizing treatment selection in Crohn’s disease: a meta-analysis of individual participant data from fifteen randomized controlled trials"

### SUPPLEMENTARY MATERIAL

TABLE OF CONTENTS:
1) Supplementary Methods (pages 2-4)
2) Supplementary Tables (pages 5-9)
3) Supplementary Figures (pages 10-14)

#### SUPPLEMENTAL METHODS

##### DATA ACCESS

In June 2019 we performed a search of clinicaltrials.gov to identify candidate studies to include in this planned meta-analysis. We identified 90 studies that were annotated as being completed, phase 2-4, randomized, double-blinded, interventional trials of treatments for Crohn's disease at the FDA-approved route, dose, and frequency. We manually confirmed 16 trials as meeting these criteria. To ensure comparability of the included cohorts and outcomes, we reviewed the major inclusion and exclusion criteria of all studies and confirmed that the Crohn's Disease Activity Index (CDAI) had been captured at week six relative to treatment initiation. We also used the Cochrane Risk of Bias 2 tool to ensure that all included studies were at a low risk of bias (Supplemental Data). Following inquiries with the sponsors of these trials, we successfully obtained access to the IPD for 15 studies (N=5703). These studies were conducted between 1999 and 2015 and corresponded to all six FDA-approved biologics as of 2019. All sponsors and data sharing partners agreed to place their data on a common, secure computing platform (Vivli) to facilitate downstream analysis.

##### QUALITY CONTROL, HARMONISATION, MISSING DATA

We performed extensive quality control evaluations of the included trials and data. This included confirming our ability to reproduce published statistics on the trial cohorts at baseline as well as the study primary endpoint. We were able to exactly reproduce most of the study results. Where discrepancies occurred, they were generally minor and fell within a 10% error bound. We reported major discrepancies to the study sponsor as per agreement. We attempted to completely eliminate all discrepancies, but this was not possible due a variety of factors, including lack of access to the original analytic code or the complete analytic dataset, and inability to contact the original analysts.

We completed an assessment of data availability for all study variables. Target variables included demographic features, CDAI at baseline and week eight, baseline inflammatory biomarkers, concomitant steroid and immunomodulator use, history of treatment with anti-TNFs, and other disease-related features. We identified nine variables that were universally available across all trials and thus could be used for downstream modelling: Age, Sex, BMI, baseline CDAI, c-reactive protein (CRP), history of TNFi use, oral steroid use, immunomodulator use, and ileal involvement.

Only 3% of the participants had at least one missing covariate at baseline. Continuous variables were addressed by median imputation, and participants with missing categorical variables were dropped from the dataset (N=86). 11% of the participants had a missing value for the outcome at week eight. To handle this, we used last-observation-carried-forward to impute these values, typically using measurements from week six and four. This is the typical practice for the analysis of these trials in regulatory submissions and was the prespecified approach in the protocols for all included trials. The variable corresponding to a history of TNFi use was available in all recent trials that occurred after the approval of the very first TNFi medication. Older trials of the first TNFis commonly excluded patients who had a history of exposure to other drugs from this class but did not include this feature as an actual variable in the data set. In these cases, we deterministically imputed this variable corresponding to no prior use.

Other variables of a priori importance could not be included in this study. Ethnicity was not collected in most trials. Race was missing in some trials, but when it was captured, it reflected significant imbalance (88% of participants were white). Other disease specific variables such as disease behavior and duration were also not uniformly captured across studies and thus could not be included in this meta-analysis.

The included trials had a range of study designs. We included both randomized and unblinded/open-label cohorts. For trials involving post-randomization gating (e.g., EXTEND, CLASSIC), we included those cohorts that were consistently exposed to a given treatment for six weeks only when post-randomization gating was not conditioned on treatment response (e.g., rerandomization of all participants, rather than just those with a particular response).

##### DRUG CLASS MODELLING, SUBGROUP IDENTIFICATION

We used sequential regression and simulation to 1) normalize all trials to a common background (placebo response), and 2) analytically isolate the portion of the patient response that could specifically be attributed to a given treatment, rather than what would have been observed without treatment (i.e., placebo; Figure 1b). For each drug class, we fit a separate linear mixed effects regression model of the drug-attributable reduction in CDAI. This outcome was modelled as a function of the nine primary variables (see the 'Quality Control' section above) handled as fixed effects, with trial as a random effect to control unmeasured heterogeneity across trials. We compared these models to intercept-only models using the likelihood ratio test. The latter corresponds to a model that ignores the role of patient-level characteristics in determining response to treatment and reflects the assumptions of methods that compare drugs based on their average effects, such as network meta-analyses. We performed Wald tests to identify significant predictors of responses to individual drug classes.

We applied the three finalized model objects to the covariate vectors of each of the 5703 participants in our meta-analysis to obtain their simulated response under each of these three counterfactual scenarios: treatment with an anti-TNF vs anti-integrin vs anti-IL-12/23. The inferred normal distributions of the conditional mean response to each drug class were pairwise compared against each other using the median of bootstrapped predictions and bootstrapped standard errors. We applied a nominal p-value threshold of 0·05 to identify patients belonging to a particular subgroup, defined as having a distinct pattern of ordinal preferences across all three drug classes. These included superiority of one drug class to another as well as indifference (lack of evidence for a difference at the p=0·05 threshold).

Because the primary focus of this study involved the testing of only three primary hypotheses (i.e., goodness of fit for each of the drug class regression models compared to intercept-only models), we used nominal p-value thresholds of 0·05 for all other hypothesis tests including the post-hoc assessments of drug subgroup membership.

SUBGROUP ASSIGNMENT

For each trial-based patient (N=5703) we predicted each drug class efficacy using the drug class models (Table 2; random effects set to 0) and estimated the 95% prediction interval using bootstrapping, the gold standard approach for deriving prediction uncertainty from linear mixed models[14]. We performed 10,000 simulations per patient. We conducted paired sample t-tests ($p < 0·05$) to further determine if any two drug class pairs were equivalent or different in efficacy to obtain a personalized treatment recommendation (Table 3). Finally, patients were assigned a subgroup based on their personalized treatment outcome based on the rank order and drug class comparisons.

DECISION SUPPORT TOOL PROTOTYPE

The decision support tool has been developed to provide real-time feedback to clinicians selecting treatments for patients with moderate-to-severe Crohn's disease. However, we are also making a prototype of the tool publicly available to enable early feedback from many potential users and to provide insights to patients wishing to understand the potential advantages and disadvantages of available treatment options.

To use the decision support tool, users must input various data points, including the patient's age, gender, body mass index (BMI), recent c-reactive protein levels (measured in milligrams per liter), current corticosteroid and immunomodulator use (yes/no), prior anti-tumor necrosis factor use (yes/no), ileal involvement (yes/no), and the Crohn's Disease Activity Index (CDAI) score. All inputs, except for the CDAI score, are mandatory for the calculation process. If any inputs are left blank, the user will receive an error message (Figure 3a) and be prompted to input a default of '0' for numeric inputs or 'No' for binary inputs if unknown. If the CDAI score is unknown, the user can either 1) leave it blank, which will result in the tool imputing a score of 300 (indicative of moderate-to-severe disease), or 2) use the MDCalc CDAI calculator[15] to obtain a precise result.

If all inputs are valid, the dashboard will output the patient's treatment recommendations in both textual and graphical forms (Figure 3b). To achieve faster recommendations in a real-time context compared to what would otherwise be obtained using bootstrapping, we used an analytical approximation for the standard error of a new prediction[16]. We used these standard errors to perform t-tests of the predicted mean response at week 6 for each pair of drug classes.

UNIVERSITY OF CALIFORNIA HEALTH DATA WAREHOUSE

The University of California (UC) Health Data Warehouse (UCHDW) contains data on 8·7 million patients who have been seen at a UC facility since 2012; data has been stored using the Observational Medical Outcomes Partnership (OMOP) data model. Additional information about the OMOP common data model can be found at https://www.ohdsi.org/data-standardization/.

We queried the UCHDW to approximate a real-world subpopulation with similar characteristics to that of the anti-IL-12/23 subgroup found in our analysis, which consists of primarily older (>50 years old) and female participants. Queries were run on April 5th, 2023. We filtered patients in the UCHDW based on diagnoses (Crohn’s disease), medication prescriptions (adalimumab, ustekinumab, infliximab, natalizumab, vedolizumab, certolizumab pegol), medication start date, current age (as of 2023), and gender (Supplemental Data). We identified standard concept ids for diagnoses and medications using the SNOMED International SNOMED CT Browser, Athena[17]. The codes are listed here: Crohn's disease (201606), adalimumab (1119119), ustekinumab (40161532), infliximab (937368), natalizumab (735843), vedolizumab (45774639), and certolizumab pegol (912263). For more details on the query, please find the code on https://github.com/rwelab/CrohnsRx.

SAMPLE SIZE CALCULATIONS

We performed simulations to calculate the expected power of a prospective trial designed to test a key prediction of our model, that anti-IL-12/23 drugs are superior to anti-TNF drugs in women over 50. In each of 1000 simulations, we sampled from the overall trial population to create pairs of study arms consisting of women over 50. Sampling was done with replacement. We used the placebo and drug-models to calculate the individual-level probability of achieving a CDAI reduction of ≥ 100 (i.e., clinical response), under an assumption of conditional normality. These were averaged within each simulated study arm, used to calculate the expected number of participants in clinical response, and then compared using a chi-squared test with an alpha of 0·05. This overall simulation procedure was performed using study arm pairs of sizes 100, 250, and 500. We repeated this analysis with the simpler inclusion criteria of just requiring participants to be over age 50, irrespective of gender.

STATISTICAL COMPUTING, WEB APPLICATION DEVELOPMENT

Programming was performed in R (version 4.2.2). We used *RShiny* to prototype a decision support tool implementing our models (https://crohnsrx.org). The analytical code was reviewed by a second member of the team and has been placed on GitHub (https://github.com/rwelab/CrohnsRx). The web dashboard utilizes manually inputted data to produce recommendations based on our models. However, for users seeking to deploy this dashboard locally, an additional mode has been made available that automatically sources input data from an OMOP-formatted EHR database.

#### SUPPLEMANTARY TABLES

| Trial | Age ≥ 18 | Baseline CDAI 220-450 | Ileal and/or colonic involvement | Disease activity by biochemical/ imaging/ endoscopy | TNFi intolerance/ failure | TNFi naive* | Stable concomitant medications | No symptomatic stricture | No abscess | No recent surgery | No stoma or ostomy |
|---|---|---|---|---|---|---|---|---|---|---|---|
| CERTIFI | ✔ | ✔ | ✔ | ✘ | ✔ | ✘ | ✔ | ✔ | ✔ | ✔ | ✔ |
| UNITI 1 | ✔ | ✔ | ✔ | ✘ | ✔ | ✘ | ✔ | ✔ | ✔ | ✔ | ✔ |
| UNITI 2 | ✔ | ✔ | ✔ | ✔ | ✘ | ✔ | ✔ | ✔ | ✔ | ✔ | ✔ |
| ENACT | ✔ | ✔ | ✔ | ✔ | ✘ | ✘ | ✔ | ✔ | ✔ | NA | ✔ |
| GEMINI 2 | ✔ | ✔ | ✔ | ✔ | ✔ | ✘ | ✔ | ✔ | ✔ | ✘ | ✔ |
| GEMINI 3 | ✔ | ✔ | ✔ | ✔ | ✘ | ✘ | ✔ | ✔ | ✔ | NA | NA |
| ACCENT 1 | ✔ | ✔ | NA | NA | ✘ | ✔ | ✔ | ✔ | ✔ | ✔ | NA |
| CLASSIC | ✔ | ✔ | NA | NA | ✘ | ✔ | ✔ | ✔ | NA | ✔ | ✔ |
| EXTEND | ✔ | ✔ | ✔ | ✔ | ✘ | ✘ | ✔ | NA | NA | NA | NA |
| NCT00291668 | ✘ | ✔ | NA | NA | ✘ | ✔ | ✔ | ✔ | NA | NA | NA |
| NCT00552058 | ✔ | ✔ | ✔ | NA | ✘ | ✔ | ✔ | ✔ | ✔ | ✔ | ✔ |
| NCT02499783 | ✔ | ✔ | ✘ | ✔ | ✘ | ✔ | ✔ | ✔ | ✘ | ✔ | ✔ |
| PRECISE 1 | ✔ | ✔ | NA | NA | ✘ | ✔ | ✔ | ✔ | ✔ | NA | ✔ |
| PRECISE 2 | ✔ | ✔ | NA | NA | ✘ | ✔ | ✔ | ✔ | ✔ | NA | ✔ |
| SONIC | ✔ | ✔ | ✔ | NA | ✘ | ✔ | ✔ | ✔ | ✔ | ✔ | ✔ |

**Supplementary Table 1: Major inclusion and exclusion criteria of studies.** If the trial protocol was not publicly available, and if the corresponding manuscript or clinical study report was silent on a given criteria, the field was annotated as NA. If the trial protocol was available and if it was clear that a given criterion was not applied for cohort selection, the field was annotated with an X. In many trials (e.g., ENACT), a history of TNF-naive or intolerance/failure was not a requirement and was captured as a participant-specific covariate for regression-based control. In other scenarios, the trial-specific covariate implicitly applied to all trial participants (e.g., TNF-naive status in PRECISE1). Trials were generally consistent on the target patients of study in terms of inclusion and exclusion criteria. To address the possibility of residual heterogeneity due to the lack of perfectly consistent eligibility criteria or unmeasured covariates, a trial-specific random effect was included in the final regression models. TNFi = tumor necrosis factor inhibitor. *Either absence of exposure or absence of prior intolerance or inadequate response.

| Drug Class Preference | Subgroup | N (%) |
|---|---|---|
| Anti-TNF,<br>Anti-Integrin | (TNF = INT) > IL | 14 (0·2) |
| Anti-TNF | TNF > IL > INT | 113 (2) |
| | TNF > INT > IL | 427 (7·5) |
| | TNF > (IL = INT) | 2,386 (42) |
| Anti-TNF, Anti-Interleukin-12/23 | (IL = TNF) > INT | 830 (14·5) |
| Anti-Interleukin-12/23 | IL > INT > TNF | 4 (0·07) |
| | IL > TNF > INT | 63 (1) |
| | IL > (TNF = INT) | 296 (5) |
| Anti-Interleukin-12/23,<br>Anti-Integrin | (TNF = INT) > IL | 2 (0·03) |
| No Preference | (TNF = IL = INT) | 1,568 (27) |

**Supplementary Table 2: Treatment Subgroups with Lower Threshold (alpha = 0·20).** The finalized mixed effects models were used to simulate counterfactual outcomes under all possible treatment scenarios. The modeled outcomes and the associated uncertainties in these outcomes were used to perform pairwise t-testing to assess evidence for rank-ordered preferences across drug classes. Distinct patterns of rank-orderings were used to establish membership in one of 9 subgroups. Subjects without sufficient statistical evidence (alpha = 0·2) of a more efficacious response to any one drug classes were placed into a 7$^{th}$ category (no preference).

| | Female (N=104) | Male (N=35) | Overall (N=139) |
|---|---|---|---|
| **Age -** Mean (SD) | 58 (± 8·4) | 63 (± 7·5) | 59 (± 8·3) |
| **BMI -** Mean (SD) | 23 (± 3·4) | 24 (± 3·0) | 23 (± 3·3) |
| **CRP -** Mean (SD) | 14 (± 17) | 13 (± 12) | 14 (± 16) |
| **History of Anti-TNF Use -** N (%) | 102 (98 %) | 32 (91 %) | 134 (96 %) |
| **Steroid Use -** N (%) | 95 (91 %) | 33 (94 %) | 128 (92 %) |
| **Immunomodulator Use -** N (%) | 18 (17 %) | 8 (23 %) | 26 (19 %) |
| **Ileal Disease -** N (%) | 80 (77 %) | 30 (86 %) | 110 (79 %) |
| **Baseline CDAI -** Mean (SD) | 280 (± 42) | 260 (± 40) | 270 (± 43) |

**Supplementary Table 3: Baseline characteristics of the anti-interleukin-12/23 subgroup, stratified by gender.** Characterization of baseline covariates of the anti-interleukin-12/23 subgroup (N = 139).

| | UC Health Data Warehouse (N = 24,012) | Clinical Trials (N = 5,703) |
|---|---|---|
| **Age (at diagnosis) - Mean (SD)** | 42 (± 19) | 38 (± 13) |
| **Gender: Female - N (%)** | 12,481 (52%) | 3,013 (53%) |
| **Race - N (%)*** | | |
| White | 15,682 (74%) | 3,320 (88%) |
| Asian | 1,161 (5%) | 292 (8%) |
| Black or African American | 1,079 (5%) | 87 (2%) |
| Other | 3,318 (16%) | 90 (2%) |
| Unknown | 2,772 (11%) | 1,914 (34%) |
| **Ethnicity - N (%)*** | | |
| Hispanic or Latino | 2,094 (10%) | 7 (1%) |
| Not Hispanic or Latino | 19,717 (90%) | 503 (99%) |
| Unknown | 2,201 (9%) | 5,193 (91%) |

**Supplementary Table 4: Demographic comparison of patients with Crohn's disease as captured by the University of California Health Data Warehouse (UCHDW), versus the meta-analyzed cohort of randomized controlled trial subjects.** The University of California (UC) Health Data Warehouse column corresponds to all patients at UC Health who received at least one diagnosis code for Crohn's Disease (ICD-9: 555.*; ICD-10: K50.*) as of June 18th, 2023. Age of diagnosis was approximated using the earliest date at which a patient received a corresponding ICD code within the UC Health system. Other race includes: Other Race (2547), Multirace (636), American Indian or Alaska Native (74), Native Hawaiian or Other Pacific Islander (61). *: To estimate the proportion of cohort subjects belonging to different racial and ethnicity categories and to facilitate comparisons across cohorts, we used the non-unknown subset as the denominator (e.g. 21,811 with non-missing race for UCHDW). Thus, the percentages within the non-unknown subsets add to 100%. This reporting approach assumes that patients with unknown race or ethnicity are representative of the overall cohort.

| Arm Size | Cohort 1: 50 years or older (N = 1040) | Cohort 2: 50 years or older, female (N = 577) | Cohort 3: 50 years or older, male (N = 463) |
|---|---|---|---|
| 100 | 0·585 | 0·755 | 0·346 |
| 250 | 0·873 | 0·969 | 0·634 |
| 500 | 0·974 | 0·999 | 0·807 |

**Supplemental Table 5: Simulation results.** We conducted a power analysis to evaluate the number of participants required per trial arm (100, 250, 500) for a hypothetical head-to-head randomized controlled trial comparing anti-IL-12/23 drugs against anti-TNF drugs for three cohorts of patients over 50 years old: men and women (cohort 1), women only (cohort 2), and men only (cohort 3). The results correspond the proportion of trials where the anti-IL-12/23 arm significantly outperformed the anti-TNF arm. We the mean over 1000 simulations for each pair of cohort and arm size.

#### SUPPLEMENTARY FIGURES

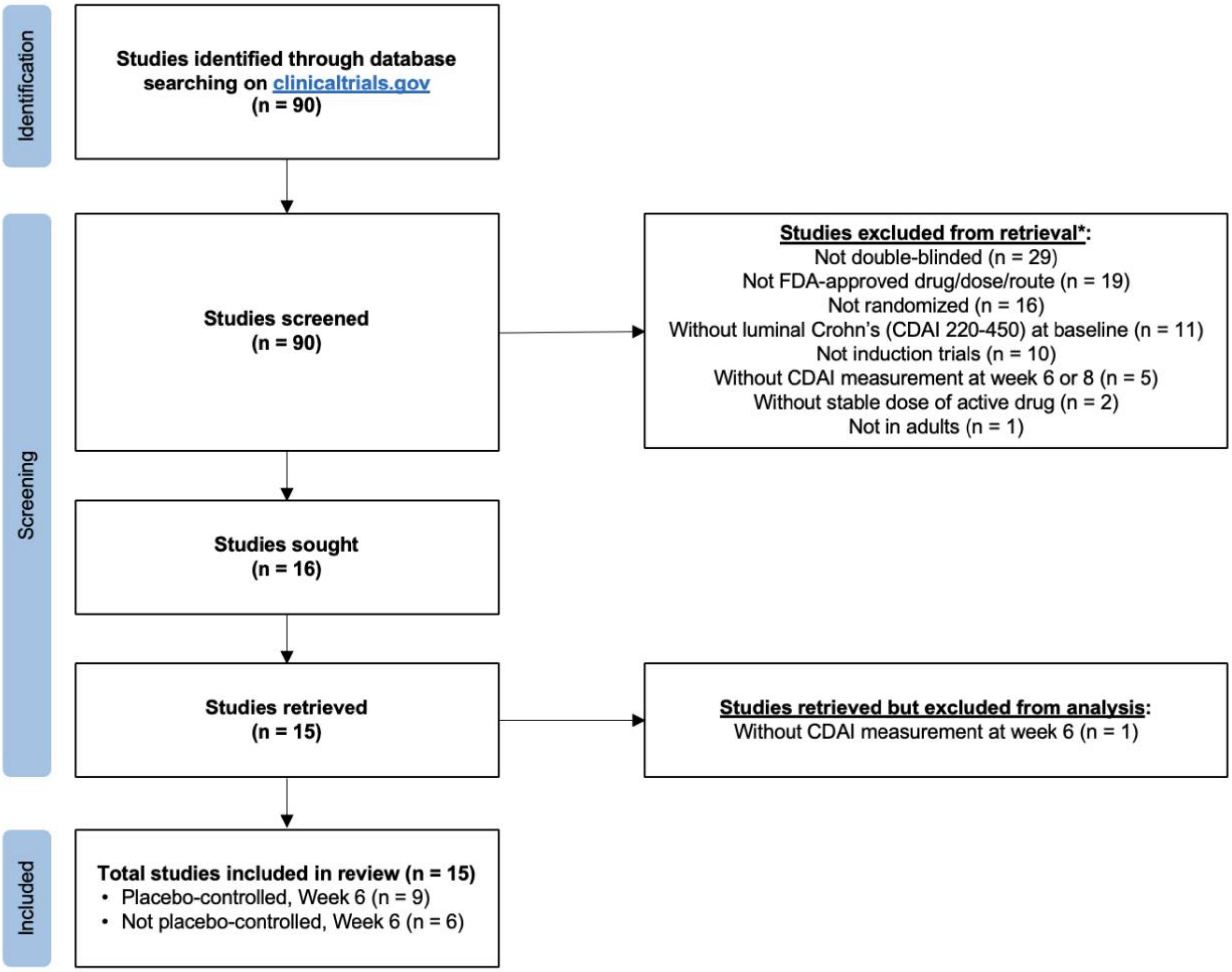


**Supplemental Figure 1: PRISMA flow diagram.** Flow diagram illustrating selection of studies.
*Some studies met more than one criterion.
†All 15 studies were retrieved and consolidated on the Vivli platform; however, only 9 studies were used for analysis as these studies captured CDAI measurement at week 8 and could be compared with the SEAVUE study.

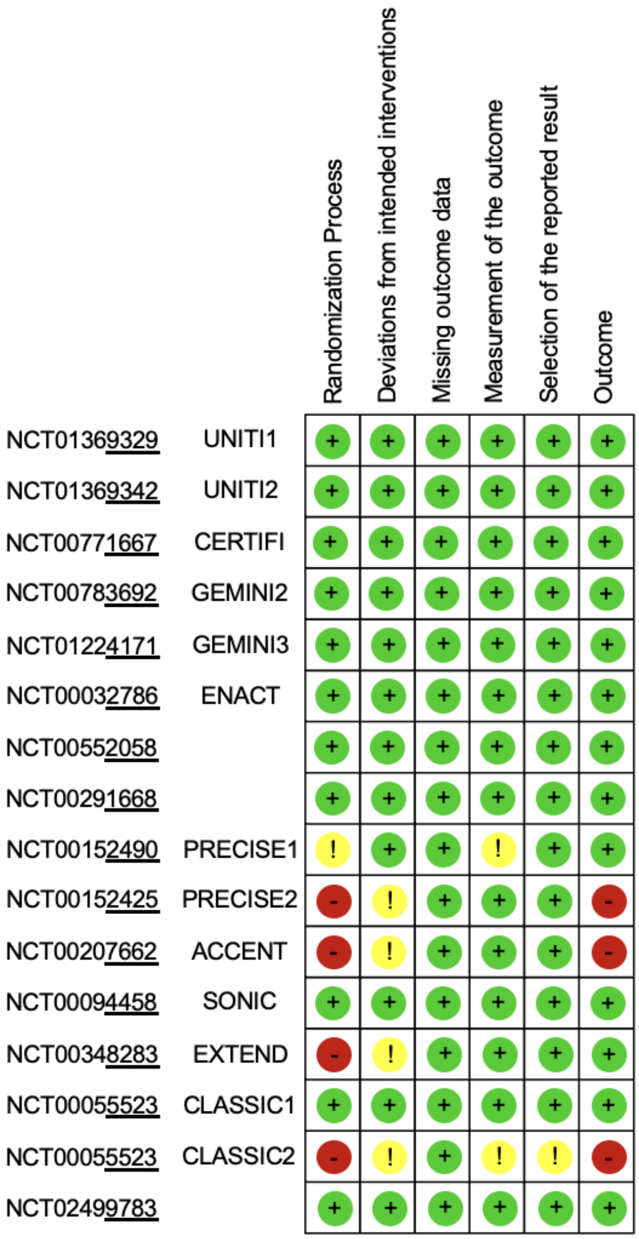


**Supplemental Figure 2: Risk of bias 2.** Cochrane's risk-of-bias tool for randomized trials version 2 (ROB2). Green, yellow, and red indicate low, moderate, and high risk of bias respectively.

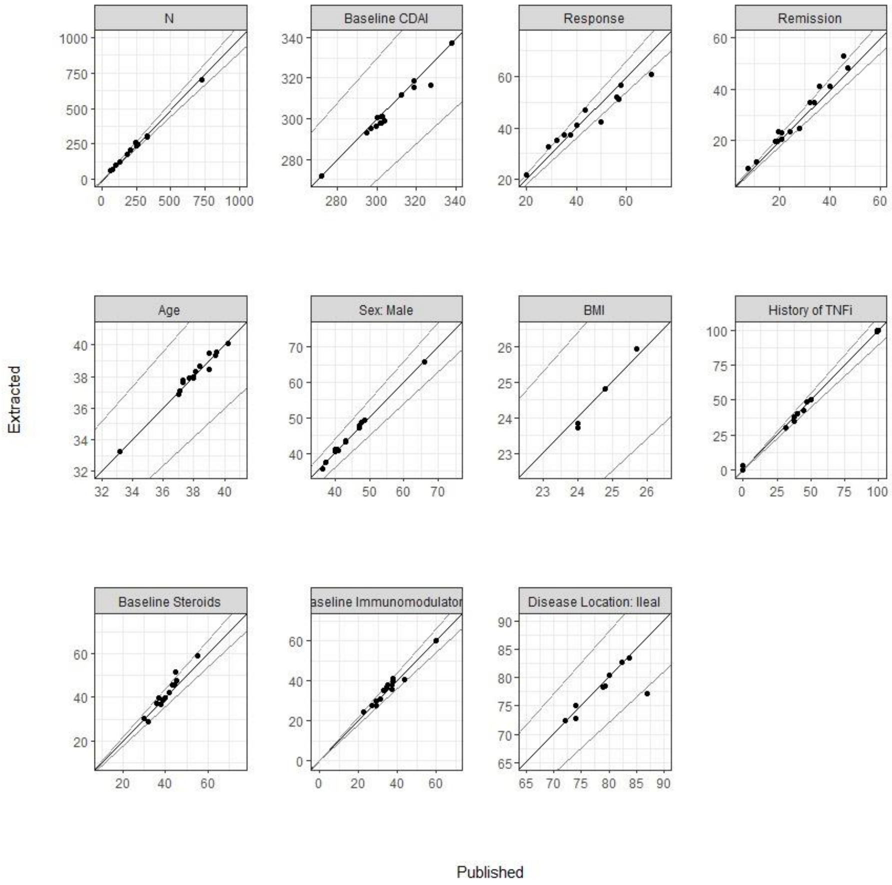


**Supplementary Figure 3: Quality Control**. Points correspond to descriptive statistics from each study as recalculated from the obtained, raw data (Y-axis) compared to the published results (X-axis).

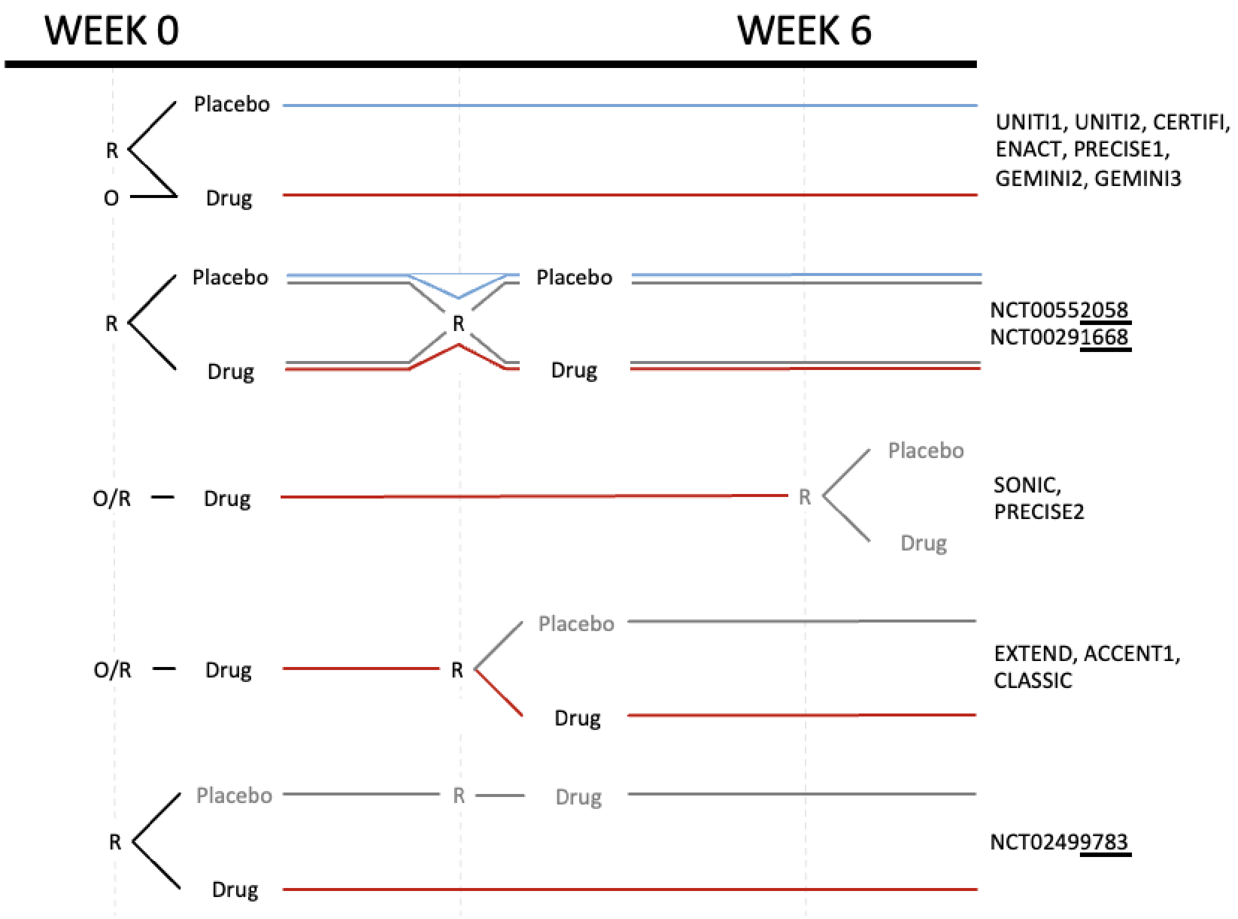


**Supplemental Figure 4: Study design summaries.** Data harmonization required careful understanding of the study designs. All treatment arms that involved 6 weeks of consistent exposure to either placebo or (blue) or active treatment at the FDA-approved doses (red) were included. R = randomized and blinded; O = open label.

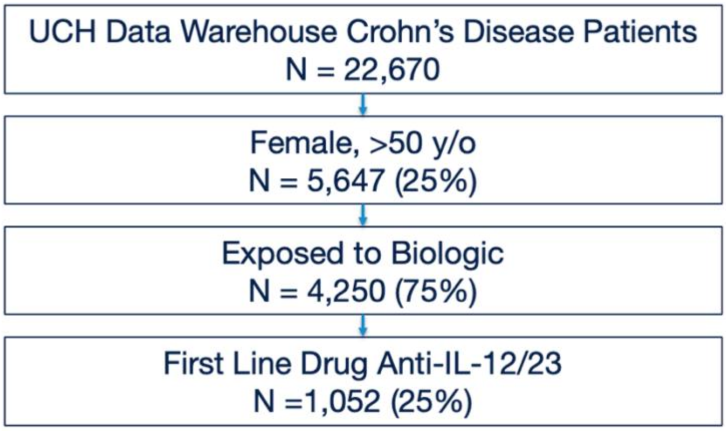


**Supplemental Figure 5: Flow diagram of estimated real-world anti-interleukin-12/23 cohort in University of California Health Data Warehouse.** Flow chart of real-world patients in the University of California Health (UCH) Data Warehouse that correspond to the anti-IL-12/23 subgroup (approximately women over 50 years of age (see Figure 2)). Of the 22,670 patients diagnosed with Crohn's disease in the database, 5647 (25%) are women over the age of 50. 4,250 of those women received their first biologic after 2016, when all biologics in the analysis were open on the market. Of those, only 1,052 (25%) women received an anti-IL-12/23 as their first line biologic. Query was run on April 5th, 2023.
